# Impact of COVID-19 pandemic on childhood immunization coverage in Indonesia: lesson learned from a nationwide analysis of the Expanded Programme on Immunization

**DOI:** 10.64898/2026.04.16.26350989

**Authors:** Amalia Nurina, Evelyn Puspaningrum, Gertrudis Tandy, Debsy Pattilima, Badriul Hegar, Grace Wangge, Raph L. Hamers, Iqbal RF Elyazar, Henry Surendra

**Affiliations:** Oxford University Clinical Research Unit Indonesia, Faculty of Medicine, Universitas Indonesia, Jakarta, Indonesia; Infectious and Tropical Diseases Epidemiology, Public Health Program, Monash University Indonesia, Tangerang, Indonesia; Directorate of Immunization, Ministry of Health of Indonesia, Jakarta, Indonesia; Indonesia Medical Education and Research Institute, Faculty of Medicine, Universitas Indonesia, Jakarta, Indonesia; Centre for Tropical Medicine and Global Health, Nuffield Department of Medicine, University of Oxford, Oxford, UK

**Keywords:** COVID-19, pandemic, immunization, vaccine preventable diseases

## Abstract

**Background:** The COVID-19 pandemic disrupted childhood immunization programmes in many countries worldwide. However, evidence on its impact in low and middle-income countries remains limited. This study examined the impact of the COVID-19 pandemic on childhood immunization coverage across 514 districts in Indonesia and identified district-level associated factors.

**Methods:** We conducted a nationwide longitudinal analysis of the Expanded Programme on Immunization to compare immunization coverage before and after the pandemic. The outcome variable was the annual childhood immunization coverage (proportion of children aged 0-12 months who have received all recommended doses of childhood immunization as per the national immunization schedule). The explanatory variables include COVID-19 burden and vaccination rates, health system and human development indicators. Mixed-effect logistic regression was done to assess association between the explanatory and outcome variables.

**Results:** At the national level, the coverage was 83.2% in pre-pandemic, 75.0% in the first year of pandemic, and 88.6%, in the second. In the first year, 69.3% of districts experienced significant decline, with a lower national coverage ratio of 0.92 (95% confidence interval 0.89–0.94). In the second year, 36.2% districts were still affected. The multivariable analysis showed that a significant decline in coverage during the first pandemic year was associated with high COVID-19 incidence (adjusted odds ratio 2.19, 95%CI 1.01–4.73 for the highest vs. lowest group), low midwife adequacy (5.84, 2.40–14.16 for the lowest vs. the highest group, 2.61, 1.26–5.40 for low-middle vs. the highest group), and a high proportion of health facility-based births (2.98, 1.49–5.98 for middle-high vs. the lowest group).

**Conclusions:** The COVID-19 pandemic negatively and unevenly impacted childhood immunization in Indonesia, with greatest impacts in districts facing a higher COVID-19 burden and weaker health system capacity. These findings underscore the need for targeted efforts to strengthen the local health system for future health crises.

**Summary box:** *What is already known on this topic:*

- According to the WHO Pulse Survey, routine immunizations were the most disrupted essential health services during the COVID-19 pandemic, reported by 70% of countries. Southeast Asia experienced the steepest drop in childhood immunization coverage compared to the other regions.
- Indonesia had the highest number of COVID-19 cases and related mortality in Southeast Asia. However, the magnitude and heterogeneity of the impact of COVID-19 pandemic on childhood immunization coverage across all 514 districts in Indonesia has not been evaluated.

*What this study adds:*

- This study affirmed that the COVID-19 pandemic greatly impacted childhood immunization coverage, disproportionately impacting district with vulnerable health systems capacity.

*How this study might affect research, practice, or policy:*

- This study highlights the critical need of addressing health inequity to strengthen health system resilience for future global health crises.
- In the context of a decentralised health system such as in Indonesia, coordination and prioritisation of available resources and public health intervention will be critical to ensure optimal health outcomes for children living in districts with weak health systems.

## Introduction

Childhood immunization remains an effective and cost-efficient public health intervention to reduce child mortality and morbidity. Maintaining high vaccination coverage is crucial for preventing vaccine preventable diseases (VPDs) outbreaks and sustaining health outcomes [1, 2]. Globally, routine vaccination has contributed to a 24% decrease of mortality rate in children under 5 years old and estimated to protect over 24 million people from poverty in 2030. Similarly in the Southeast Asia region, immunization programs have successfully eliminated transmission of wild-type polio and reduce prevalence of measles, Japanese encephalitis, and hepatitis B [3]. In response to the global success, the Global Immunization Agenda 2021 – 2030 was launched to avert 50 million deaths, eradicate polio, reduced VPDs outbreaks, and promote equitable access by reducing half proportion of unvaccinated children and increase vaccination coverage up to 90% [2]. However, progress in 2023 revealed an estimation of 14.5 million of children under 1 year old remaining unprotected by basic vaccination (*zero dose* children), with 75% of this concentrated in 14 countries including Indonesia [4].

The COVID-19 pandemic, caused by SARS-CoV-2 virus, severely disrupted immunization progress towards the global target. According to the WHO Pulse Survey in 2020, routine immunization and outreach services were the most frequently impacted health services during the pandemic [5]. Global basic immunization coverage experienced a decline from 86% in 2019 to 83% and 81% in 2020 and 2021, respectively. The Southeast Asia region has the steepest decline compared to other regions with 91% coverage in 2019 to 83% in 2021 [6, 7]. The WHO and GAVI (the Vaccine Alliance) reported that 66% of GAVI-eligible countries experienced vaccination program disruption due to shipment delays and vaccine stockouts [8].

Indonesia, home to over 280 million population, ranked as the fourth most populated country characterized by its high socioeconomic, geographical, and cultural diversity across the nation [9]. In 2020, health program planning and budgeting responsibilities have been decentralized to 514 administrative districts, resulting in vast heterogeneity of health measures and unequal access to health services across the archipelago [10]. The country’s Public Health Development Index (PHDI) in 2018 reflected these disparities, with scores ranging from 35% in Paniai district, a low resource setting located in Central Papua province, to 74.7% in Gianyar district, a higher resource setting located in Bali province [11].

The COVID-19 pandemic further exposed the vulnerability of Indonesia’s pre-existing health system gap [12]. During the pandemic, the country experienced the highest number of COVID-19 cases and mortality in the Southeast Asia region. Following the first reported case in March 2020, the government decided to restrict mobilization at the national level, which caused the national immunization program to be suspended. As a result, Indonesia ranked among the top three countries with the highest number of *zero dose children* in 2021, leaving over one million children unprotected [1]. By 2023, the impact became more evident, Indonesia placed among the top ten countries with large-scale destructive measles outbreaks, while at the same time grappling with vaccine-derived polio outbreaks in Aceh and West Java province [13, 14].

This study aimed to assess the impact of COVID-19 on Indonesia’s childhood immunization coverage across its 514 districts by comparing the pre-pandemic, pandemic and post-pandemic years. We also explored district-level factors, including COVID-19 burden and vaccination coverage, and indicators of human development and health systems capacity associated with the COVID-19 impact on immunization coverage. The findings will inform evidence-based insights to strengthening health systems resilience for future public health emergencies.

## Methods

### Study design

We conducted a nationwide longitudinal analysis of the government Expanded Programme on Immunization from January 2019 to December 2024. District-level data were obtained from the Ministry of Health immunization program covering 514 districts across 34 provinces in Indonesia from 2019 to 2021. Since the first COVID-19 case was reported in March 2020, we defined our study timeframe as pre-pandemic period (January 2019 to February 2020), first year of pandemic (March 2020 to February 2021) and second year of pandemic (March 2021 to December 2021). To illustrate the post pandemic trend, we collected publicly available province-level data on annual vaccine coverage from Indonesia’s annual health profile report (2022-2024).

### Patient and public involvement

This study did not include patients and public in the design, or conduct, or reporting, or dissemination plans. This study was a secondary analysis of anonymised aggregated routine programme data.

### Data collection

The main outcome of interest was annual childhood immunization coverage. The Indonesian Ministry of Health defines childhood immunization coverage as a proportion of children aged 0-12 months who have received all recommended doses of childhood immunization as per the national immunization schedule [15]. The immunization includes 1 dose of Hepatitis B vaccine, 1 dose of BCG vaccine, 3 doses of DPT-HB-HiB vaccine, 4 doses Polio Vaccine, and 1 dose of measles-containing vaccinee [16].

To measure COVID-19 burden, we collected district-level data on the number of COVID-19 cases and related mortality from March 2020 until December 2021. Data were extracted from the National COVID-19 Task Force database [17]. We also included data on the district-level COVID-19 vaccination coverage from the National COVID-19 vaccination database (January 2021 to March 2022), as a proxy of additional burden to the health system during the pandemic crisis [18]. To analyse the district-level public health development, we utilized the latest available data from the 2018 Public Health Development Index [11], specifically reflecting child health development and health services capacity. These include the proportion of stunting and malnutrition, coverage of child weight monitoring and neonatal visit, proportion of mothers who give birth in health facilities (health facility birth), proportion of sub-district with adequate doctor, proportion of village with adequate midwives, proportion of village with adequate integrated community-based health post (*posyandu)*, and national insurance ownership. To ascertain human development and sociodemographic factors, we obtained district-level data on per-capita domestic expenditure, life expectancy at birth, and average length of basic formal education from the 2020 Human Development Index report [19] and the annual population number from January 2019 to December 2021 [20]. Operational definitions and data source of all included variables were described in table 1.

**Table 1.** List of variables included in the study.

| No | Variables | Definition | Source |
| --- | --- | --- | --- |
| 1 | Childhood immunization coverage | Proportion of children who completed basic immunization according to the MoH definition by one year target (27) | Ministry of Health immunization registry |
| 2 | Cumulative COVID-19 incidence rate | Proportion of new cases of COVID-19 per 100,000 population | National COVID-19 task force |
| 3 | Cumulative COVID-19 mortality rate | Proportion of COVID-19 related mortality per 100,000 population |  |
| 4 | COVID-19 vaccination coverage | Proportion of population vaccinated against COVID-19 at first, second, and third dose. | National COVID-19 vaccine database |
| 5 | Health facility birth | Proportion of mother who gives birth in the health facility | 2018 PHDI report |
| 6 | Proportion of sub-district with adequate doctors | Proportion of sub-district in a district that equipped with adequate doctors (1:2500 population) |  |
| 7 | Proportion of village with adequate midwives | Proportion of village in a district that equipped with adequate midwives (1:1000 population) |  |
| 8 | Proportion of village with adequate integrated community-based health post (*posyandu*) | Proportion of village in a district that equipped with adequate *posyandu* (1:4) |  |
| 9 | National health insurance ownership | Proportion of population owning at least one insurance |  |
| 10 | Proportion of malnutrition children | Proportion of children under five based on weight-for-age having Z-score less than -2SD |  |
| 11 | Proportion of stunting | Proportion of children under five based on height-for-age having Z-score less than -2SD |  |
| 12 | Coverage of child weight monitoring | Proportion of children under five who were weighted in the past 12 months |  |
| 13 | Coverage of neonatal visit | Proportion of children under five who received health services within 6 to 48 hours after birth |  |
| 14 | Domestic expenditure per capita | Average annual household expenditure per person | 2020 HDI report |
| 15 | Average duration of formal education | Average number of years of schooling received by individuals under 25 years old |  |
| 16 | Life expectancy at birth | Average number of years a newborn is expected to live |  |

### Statistical Analysis

To address temporal bias in our main analysis, we divided the childhood immunization coverage into three periods: the pre-pandemic period (January 2019 to February 2020), first year of pandemic (March 2020 to February 2021), and second year of pandemic (March 2021 to December 2021). For descriptive analysis, we summarise and compare the immunization coverage at national level and by island in 2019-2021. Due to limited access to the similar district-level immunization coverage data, we could only describe a national trend covering period of 2022 to 2024, based on the report available from Indonesia Health Profile published by the Indonesian Ministry of Health. The immunization coverage was calculated for each district and period by dividing the number of children who completed childhood immunization with the number of targeted children at the same period. To assess district-level change in immunization coverage, we quantified coverage ratio by comparing immunization coverage in each pandemic year against the pre-pandemic data. A binary variable was created to classify districts as affected (1) and not affected (0). Affected districts were defined as districts with statistically significant decrease in immunization coverage (coverage ratio <1 and p<0.05). A geographical map was created to visualize the spatial heterogeneity the immunization coverage and coverage ratio during pandemic periods.

The district-level explanatory variables were transformed into quartiles (lowest, low-middle, middle-high, and highest) using quantile-based binning. We conducted bivariable and multivariable mixed-effect logistic regression to analyse the association between explanatory variables with the coverage decline during the first pandemic year and expressed as odds ratio (OR) with its respective 95% confidence intervals. Province was treated as the random-effect variable to adjust for clustering of observations within provinces. A null model analysis, wherein no predictor was added, revealed that 23.4% of the outcome variance was attributed to a clustering effect at the provincial level. This supports the necessity of mixed-effect modelling, as also confirmed by the likelihood ratio test (p<0.001). All independent variables with a p<0.20 in the bivariable analysis were considered for inclusion in the multivariable analysis. We used a forward selection approach to fit the final regression model. We set statistical significance at two-sided p<0.05. All analyses were done in Stata ver. 17. This study is reported as per Strengthening the Reporting of Observational Studies in Epidemiology (STROBE) guidelines [21].

### Ethics

This study was approved by the Health Research Ethics Committee of the National Institute of Health Research and Development, Ministry of Health of Indonesia (LB.02.01/2/KE.486/2021). The requirement for patient consent was waived as this was a secondary analysis of aggregated routine programme data with no personal identifiers.

## Results

Table 2 summarise the comparison of childhood immunization coverage over time and by island in Indonesia. At national level, the childhood immunization coverage was 83.2% in the pre-pandemic period, with the highest coverage observed in Java Island and the lowest in Maluku and Papua Islands (table 2 and online supplemental figure 1). During this period, 205 of all 514 districts met the WHO target of 90% coverage. In the first year of pandemic, the national coverage declined to 75%, leading to only 126 districts met the WHO target. The largest decrease recorded in Java Island where coverage dropped by 13.6% compared to the pre-pandemic year. In the second year of pandemic, the national coverage showed noticeable recovery reaching 88.6%, with the largest increase observed in Sulawesi Island where coverage rose by 32.3% from the previous year. The number of districts meeting the WHO target also increased by 53.5% compared to the previous year, accounting for 271 districts. Due to limited data for the post-pandemic year, similar analysis comparing number of districts meeting the WHO target could not be done. However, the official data published by the Ministry of Health suggest a significant decline, from 78.4% districts meeting national target (80% coverage) in 2022, to 71.8% in 2023, and 47.8% in 2024.

**Table 2.**
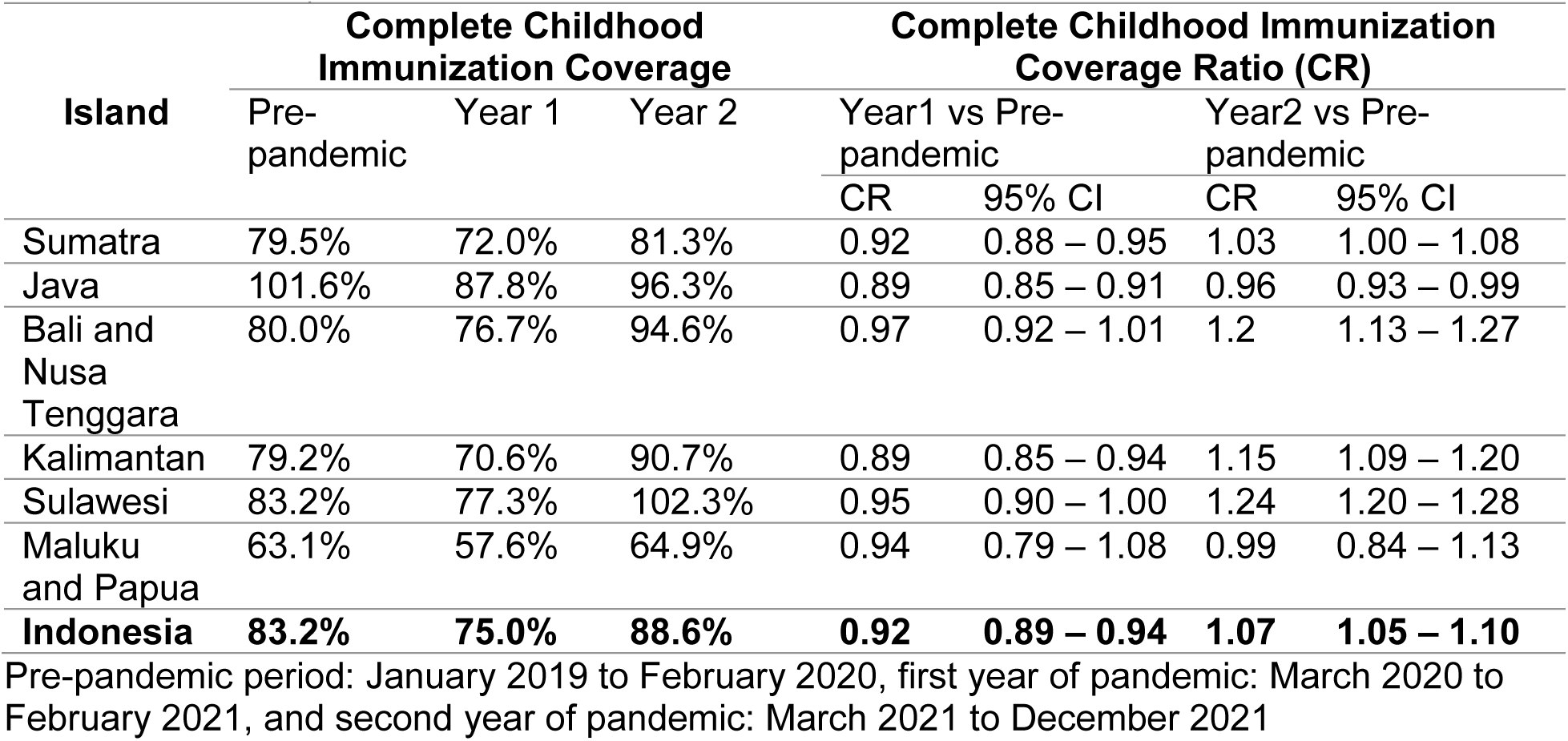
Childhood immunization coverage in the pre-pandemic (January 2019 to February 2020), first year (March 2020 to February 2021) and second year of pandemic (March 2021 to December 2021) in Indonesia.

| Island | Complete Childhood Immunization Coverage |  |  | Complete Childhood Immunization Coverage Ratio (CR) |  |  |  |
| --- | --- | --- | --- | --- | --- | --- | --- |
|  | Pre-pandemic | Year 1 | Year 2 | Year1 vs Pre-pandemic |  | Year2 vs Pre-pandemic |  |
|  |  |  |  | CR | 95% CI | CR | 95% CI |
| Sumatra | 79.5% | 72.0% | 81.3% | 0.92 | 0.88 – 0.95 | 1.03 | 1.00 – 1.08 |
| Java | 101.6% | 87.8% | 96.3% | 0.89 | 0.85 – 0.91 | 0.96 | 0.93 – 0.99 |
| Bali and Nusa Tenggara | 80.0% | 76.7% | 94.6% | 0.97 | 0.92 – 1.01 | 1.2 | 1.13 – 1.27 |
| Kalimantan | 79.2% | 70.6% | 90.7% | 0.89 | 0.85 – 0.94 | 1.15 | 1.09 – 1.20 |
| Sulawesi | 83.2% | 77.3% | 102.3% | 0.95 | 0.90 – 1.00 | 1.24 | 1.20 – 1.28 |
| Maluku and Papua | 63.1% | 57.6% | 64.9% | 0.94 | 0.79 – 1.08 | 0.99 | 0.84 – 1.13 |
| **Indonesia** | **83.2%** | **75.0%** | **88.6%** | **0.92** | **0.89 – 0.94** | **1.07** | **1.05 – 1.10** |

Pre-pandemic period: January 2019 to February 2020, first year of pandemic: March 2020 to February 2021, and second year of pandemic: March 2021 to December 2021

The impact of COVID-19 pandemic on immunization coverage varied widely across districts (figure 1 and online supplemental figure 1). In the first year of pandemic, 356 districts (69.3%) experienced a significant decline in the coverage. The national coverage ratio was 0.92 (95% CI 0.89–0.94), widely varied from 0.07 in South Monokwari district, West Papua province to 4.52 in Paniai district, Papua province (figure 1A). In the second year of pandemic, the number of significantly affected districts decreased to 186 (36,2%) with a national coverage ratio of 1.07 (95%CI 1.05–1.10), signalling a net recovery. During this period, the coverage ratio ranged from 0.0 in Pegunungan Arfak district, West Papua province to 3.86 in Paniai district, Papua province (figure 1B).

**Figure 1.**
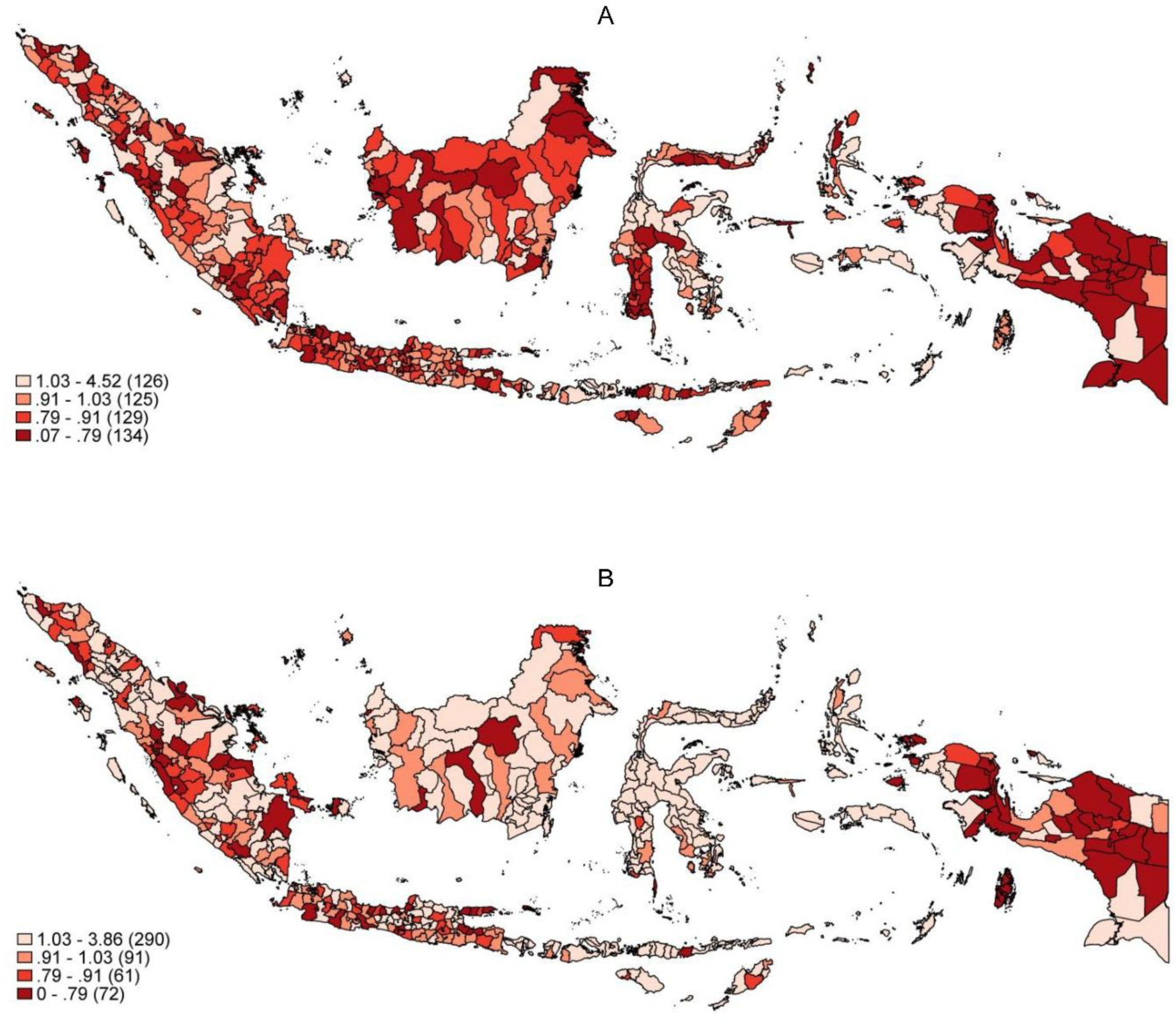
Geographical map of childhood immunization coverage ratio across Indonesia in the first year (A) and the second year of pandemic (B) against the pre-pandemic period.

Summary statistics of explanatory variables being assessed in this study is presented in online supplemental table 1. Briefly, the data suggest the heterogeneity of COVID-19 burden and vaccination coverage, PHDI, and HDI scores across the 514 districts in Indonesia.

Bivariable analysis (table 3) suggest significant association between decline in immunization coverage in the first year of pandemic with COVID-19 incidence and mortality rate, proportion of malnutrition and stunting, coverage of neonatal visit, proportion of health facility-based birth, adequacy of doctors, midwives, and *posyandu*, proportion of national health insurance ownership, domestic expenditure, average duration of formal education, life expectancy, and first dose of COVID-19 vaccine coverage. On the other hand, coverage of childhood monitoring and second dose of COVID-19 vaccine coverage was not significantly associated with the outcome, therefore were excluded in the multivariable analysis.

**Table 3.** Bivariable analysis of factors associated with decline of childhood immunization coverage in Year 1 of COVID-19 pandemic in Indonesia.

| Variables | Odds Ratio | 95% Confidence Interval | p value |
| --- | --- | --- | --- |
| **Cumulative COVID-19 incidence rate** |  |  |  |
| Lowest | 1 |  |  |
| Low-middle | 1.416 | 0.759 - 2.642 | 0.274 |
| **Middle-high** | **2.0305** | **1.045 - 3.947** | **0.037** |
| **Highest** | **3.469** | **1.686 - 7.140** | **0.001** |
| **Cumulative COVID-19 mortality rate** |  |  |  |
| Lowest | 1 |  |  |
| Low-middle | 1.565 | 0.801 - 3.059 | 0.19 |
| Middle-high | 1.607 | 0.811 - 3.183 | 0.174 |
| **Highest** | **3.631** | **1.673 - 7.881** | **0.001** |
| **Proportion of malnutrition** |  |  |  |
| Lowest | 1.891 | 0.969 - 3.692 | 0.062 |
| **Low-middle** | **2.568** | **1.348 - 4.892** | **0.004** |
| Middle-high | 1.752 | 0.973 - 3.155 | 0.062 |
| Highest | 1 |  |  |
| **Proportion of stunting** |  |  |  |
| **Lowest** | **3.095** | **1.580 - 6.063** | **0.001** |
| Low-middle | 1.357 | 0.737 - 2.494 | 0.327 |
| Middle-high | 1.305 | 0.720 - 2.366 | 0.380 |
| Highest | 1 |  |  |
| **Coverage of child weight monitoring** |  |  |  |
| Lowest | 0.738 | 0.337 - 1.616 | 0.447 |
| Low-middle | 1.519 | 0.727 - 3.173 | 0.266 |
| Middle-high | 1.522 | 0.766 - 3.025 | 0.230 |
| Highest | 1 |  |  |
| **Coverage of neonatal visit** |  |  |  |
| Lowest | 1 |  |  |
| Low-middle | 1.893 | 0.997 - 3.593 | 0.051 |
| Middle-high | 1.872 | 0.958 - 3.660 | 0.067 |
| **Highest** | **2.035** | **1.018 - 4.068** | **0.045** |
| **Proportion of health facility birth** |  |  |  |
| Lowest | 1 |  |  |
| **Low-middle** | **2.097** | **1.168 - 3.767** | **0.013** |
| **Middle-high** | **4.321** | **2.224 - 8.396** | **0.000** |
| **Highest** | **3.343** | **1.663 - 6.721** | **0.001** |
| **Proportion of sub-district with adequate doctors** |  |  |  |
| Lowest | 1 |  |  |
| Low-middle | 1.268 | 0.685 - 2.348 | 0.450 |
| **Middle-high** | **2.285** | **1.242 - 4.207** | **0.008** |
| **Highest** | **2.305** | **1.208 - 4.399** | **0.011** |
| **Proportion of village with adequate midwives** |  |  |  |
| **Lowest** | **7.026** | **3.103 - 15.906** | **<0.001** |
| **Low-middle** | **3.113** | **1.572 - 6.164** | **0.001** |
| Middle-high | 1.463 | 0.825 - 2.592 | 0.193 |
| Highest | 1 |  |  |
| **Proportion of village with adequate *posyandu*** |  |  |  |
| Lowest | 1 |  |  |
| Low-middle | 1.337 | 0.729 - 2.452 | 0.348 |
| **Middle-high** | **2.896** | **1.377 - 6.091** | **0.005** |
| **Highest** | **6.113** | **2.554 - 14.628** | **0.000** |
| **Health insurance ownership** |  |  |  |
| Lowest | 1 |  |  |
| Low-middle | 1.392 | 0.780 - 2.483 | 0.263 |
| **Middle-high** | **2.365** | **1.271 - 4.401** | **0.007** |
| **Highest** | **2.379** | **1.221 - 4.633** | **0.011** |
| **Domestic expenditure per capita (USD)** |  |  |  |
| Lowest | 1 |  |  |
| Low-middle | 1.093 | 0.580 - 2.059 | 0.783 |
| Middle-high | 1.487 | 0.765 - 2.888 | 0.242 |
| **Highest** | **2.917** | **1.425 - 5.970** | **0.003** |
| **Average duration of formal education** |  |  |  |
| Lowest | 1 |  |  |
| Low-middle | 1.033 | 0.559 - 1.911 | 0.917 |
| Middle-high | 1.553 | 0.810 - 2.975 | 0.184 |
| **Highest** | **2.623** | **1.326 - 5.190** | **0.006** |
| **Life expectancy at birth** |  |  |  |
| Lowest | 1 |  |  |
| Low-middle | 1.828 | 0.994 - 3.360 | 0.052 |
| **Middle-high** | **2.634** | **1.353 - 5.130** | **0.004** |
| **Highest** | **3.732** | **1.721 - 8.092** | **0.001** |
| **COVID-19 vaccine coverage (1<sup>st</sup> dose)** |  |  |  |
| Lowest | 2.179 | 0.983 - 4.827 | 0.055 |
| **Low-middle** | **2.506** | **1.226 - 5.121** | **0.012** |
| Middle-high | 1.596 | 0.841 - 3.028 | 0.152 |
| Highest | 1 |  |  |
| **COVID-19 vaccine coverage (2<sup>nd</sup> dose)** |  |  |  |
| Lowest | 1.107 | 0.500 - 2.451 | 0.801 |
| Low-middle | 1.372 | 0.674 - 2.791 | 0.383 |
| Middle-high | 1.029 | 0.527 - 2.011 | 0.933 |
| Highest | 1 |  |  |

Table shows results of bivariable mixed-effect logistic regression to assess the association between explanatory variables with the childhood immunization coverage decline during the first pandemic year. Province was treated as the random-effect variable.

In the multivariable analysis (table 4), a decline in coverage at the district-level was significantly associated with high COVID-19 incidence (adjusted odds ratio 2.19, 95% CI 1.01–4.73 for the highest vs. lowest group), low midwife adequacy (5.84, 2.40–14.16 for the lowest vs. the highest group, 2.61, 1.26–5.40 for low-middle vs. the highest group), and having a higher proportion of health facility-based births (2.98, 1.49–5.98 for middle-high vs. the lowest group). There was no association for COVID-19 mortality rate, proportion of malnutrition, proportion of stunting, coverage of child weight monitoring, coverage of neonatal visit, adequacy of doctors and *posyandu*, proportion of health insurance ownership, domestic expenditure, average duration of formal education, life expectancy, and COVID-19 vaccine coverage.

**Table 4.** Multivariable analysis of factors associated with decline of childhood immunization coverage in Year 1 of COVID-19 pandemic in Indonesia.

| Variable | Adjusted odds ratio | 95% Confidence Interval | P value |
| --- | --- | --- | --- |
| **Cumulative COVID-19 incidence rate** |  |  |  |
| Lowest | 1 |  |  |
| Low-middle | 1.405 | 0.739 - 2.669 | 0.300 |
| Middle-high | 1.630 | 0.814 - 3.264 | 0.168 |
| **Highest** | **2.186** | **1.010 - 4.728** | **0.047** |
| **Proportion of sub-districts with adequate midwives** |  |  |  |
| **Lowest** | **5.840** | **2.408 - 14.164** | **<0.001** |
| **Low-middle** | **2.612** | **1.264 - 5.397** | **0.010** |
| Middle-high | 1.404 | 0.779 - 2.531 | 0.259 |
| Highest | 1 |  |  |
| **Proportion of health facility birth** |  |  |  |
| Lowest | 1 |  |  |
| Low-middle | 1.772 | 0.964 - 3.259 | 0.066 |
| **Middle-high** | **2.981** | **1.486 - 5.980** | **0.002** |
| Highest | 1.441 | 0.645 - 3.218 | 0.373 |

Table shows results of multivariable mixed-effect logistic regression to assess the association between explanatory variables with the childhood immunization coverage decline during the first pandemic year. Province was treated as the random-effect variable.

## Discussion

This study explores the impact of COVID-19 pandemic on childhood immunization coverage in Indonesia, focusing on trends, geographical variation, and association with underlying district-level factors. Consistent with the Ministry of Health report [22], our analysis identified a nationwide decline in childhood immunization coverage by 8% in the first year of the COIVD-19 pandemic. Compared to the global average decline (3.5% in 2020), and regional decline in Southeast Asia (5.5%), and European regions (1%), Indonesia experienced a greater decline in immunization coverage during the first pandemic year [6]. This underscores how COVID-19 pandemic imposed significant indirect health threats beyond its related mortality and morbidity, by disrupting both supply and demand side of immunization service worldwide [23, 24]. The greater disruption of pandemic on childhood immunization was observed in another LMIC, Mexico (36% decline in 2020) [25].

In the second year of pandemic, signs of recovery of the childhood immunization emerged. The proportion of affected districts improved to 36.2% of districts within this period. In the same period, there was a gradual transition from large-scale social restriction (PSBB) to community-scale activities restriction (PPKM) which led to more mobility and reactivation of vaccine programmes. The transition allowed partial reopening of *posyandu*, where most maternal and child health services including immunization took place. However, the coverage did not rebound to the pre-pandemic level, particularly in Java Island (the most populated island in Indonesia). This added burden to the local health system and exposed healthcare workers with higher risk of COVID-19 infection and mortalities [26, 27]. Similar finding was reported in Armenia, where routine vaccination coverage remained lower than before the pandemic, despite signs of recovery [28]. Beyond the pandemic period, report from Indonesian government suggested that Indonesia continues to experience declining trend in immunization coverage from 78.4% districts meeting national target (80% coverage) in 2022, to 71.8% in 2023, and 47.8% in 2024. A recent report indicated that the expanded decline may contribute to the recent VPDs outbreak occurring in the country, placing Indonesia among the top ten countries with large-scale destructive measles outbreaks [13].

Our study identified widespread and heterogeneous impact of COVID-19 pandemic across districts, with 69.3% of districts experiencing lower immunization coverage and coverage ratio ranging widely from 0.07 to 4.52 during the first year of pandemic. This variation is likely attributed to Indonesia’s decentralized health system, where local governments have the authority to manage health budget, resources, and delivery, leading to unequal service quality and capacity across the nation. During the pandemic crises, this fragmentation contributed to heterogenous immunization performance outcome [12]. The resulting disruptions placed the country at higher risk of VPDs outbreak in children. A previous study reported that in 2021, the disruption contributed to over 1 million children missed childhood basic vaccination, leading to 415 districts receiving high risk status of polio outbreak in the following year [29].

Even without the pandemic burden, immunization coverage is influenced by complex interplay of physical (e.g. vaccine cost, supply and logistic), attitudinal (e.g. risk perception), and contextual factors (e.g. socioeconomic status, community-level health system, geographical area). Given this complexity, the uneven impact of COVID-19 pandemic on childhood immunization coverage is partially shaped by underlying district-level disparities [30, 31]. Our multivariable model helps to explain this heterogeneous impact between districts, suggesting that a high COVID-19 incidence rate, low midwife adequacy, and higher health facility-based births as strong explanatory variables of the decline in the coverage at the district level.

Although previous evidence shows that basic immunization is mainly administered in integrated community-health post or *posyandu* [22], our study found no association between *posyandu* adequacy and coverage decline at district level. This was potentially due to 75% of *posyandu* services being halted in the country and immunization were only given by appointment with the healthcare personnel [29]. Nonetheless, our finding suggests a significant association between low midwife adequacy and the coverage decline, consistent with various findings from European and Southeast Asia countries reporting similar disruption in routine immunization programs due to inadequate workforces [9]. In Indonesia, midwives play a central role as vaccinators in *posyandu*, as well as conducting health promotion and outreach. Their re-disposition to pandemic-related tasks exacerbates the disruption in districts with already limited midwife availability [32].

Interestingly, our model suggests districts with higher proportion of health facility-based birth associated with the decline in childhood immunization coverage, contradicting previous findings in Ethiopia and Nigeria [33, 34]. This discrepancy is likely explained by policy changes during the early pandemic in Indonesia. The Indonesian Ministry of Health mandated all mothers to have negative PCR test results prior to delivery in health facilities. Combined with healthcare worker shortages and re-assignment to pandemic management, facility-based delivery was simultaneously reduced by over 50% at the national level during this period [32].

Our findings have important implications for strengthening the country’s health system resilience, especially on immunization programs. The results emphasize the need to maintain healthcare workforce availability and continue maternal and child health services during future health emergencies – especially in districts with weaker health capacity, higher disease transmission, and interrupted maternal care services. Several measures include redistribution of healthcare workforces within the province, recruitment of trained volunteers, improving workload management, establishing clear policy to protect healthcare personnels and patients, and providing mental health support for healthcare personnel [27]. In addition, inclusion of non-physician vaccinators such as dentists, paramedics, and community pharmacists serves as potential long-term strategies for future pandemic preparedness, as applied in various high-income settings including the United States, United Kingdom and New Zealand [35].

Our study has several limitations. First, it did not capture all health system factors that may be important determinants of childhood immunization coverage, particularly those related to vaccine availability (e.g. cold-chain adequacy, vaccine stocks, etc.). The absence of these variables could affect the estimated association and limit the comprehensiveness of the findings. Second, our analysis only covered the first two years of the COVID-19 pandemic (until December 2021), while mobility restriction policies still prevailed. Therefore, post-pandemic recovery of immunization service is not reflected, since mobility restriction was gradually phased out at the end of 2022 [36]. Third, the ecological nature of the study design is meant to assess associations at district-level, therefore findings from this study should not be interpreted for individual-level associations. Finally, due to limited access to data, our analysis could not assess the impact of COVID-19 pandemic on the coverage of each vaccine, i.e., Hepatitis B, BCG, DPT-HB-HiB, Polio, and measles. Nonetheless, to our knowledge, our study is one of the first studies to provide essential evidence on the impact of COVID-pandemic on the combined childhood immunization programme in Indonesia.

In conclusion, the COVID-19 pandemic disrupted the childhood immunization programme in Indonesia, leading to the decline in coverage, which extended beyond pandemic period. This study provides key lessons for strengthening the country’s health system resilience especially in maternal and child health services in preparation for future health crises. Further exploration is needed to assess factors associated with the prolonged decline in the childhood immunization coverage in the post-pandemic period. Future study evaluating trend and factors associated with vaccine-specific coverage is needed.

## Contributors

IRFE and RLH were the principal investigators. AN, HS, IRFE and RLH conceptualized this study. EP, HS and IRFE collected and verified the data. AN and EP conducted analysis and prepared the manuscript with supervision from HS. AN, GW HS, IRFE drafted the manuscript. All authors reviewed the full manuscript and approved the content of the final manuscript for submission purpose. All authors had access to the full dataset used in this study.

## Data Availability Statement

Following publication, the dataset used in this study will be made available with reasonable request via email to the corresponding author, including a detailed research proposal, study objectives, and statistical analysis plan.

## Acknowledgement

We express our gratitude to the sub-directorate of immunization, Ministry of Health Indonesia and the district health for granting access to the national immunization registry. We also thank all collaborators in the COHERE project, in particular Ministry of Health of Indonesia for the immense contribution to this study. We acknowledge all healthcare workers involved in the national immunization program and field data collection.

