## supplemental for "Impact of COVID-19 pandemic on childhood immunization coverage in Indonesia: lesson learned from a nationwide analysis of the Expanded Programme on Immunization"

### Supplemental material

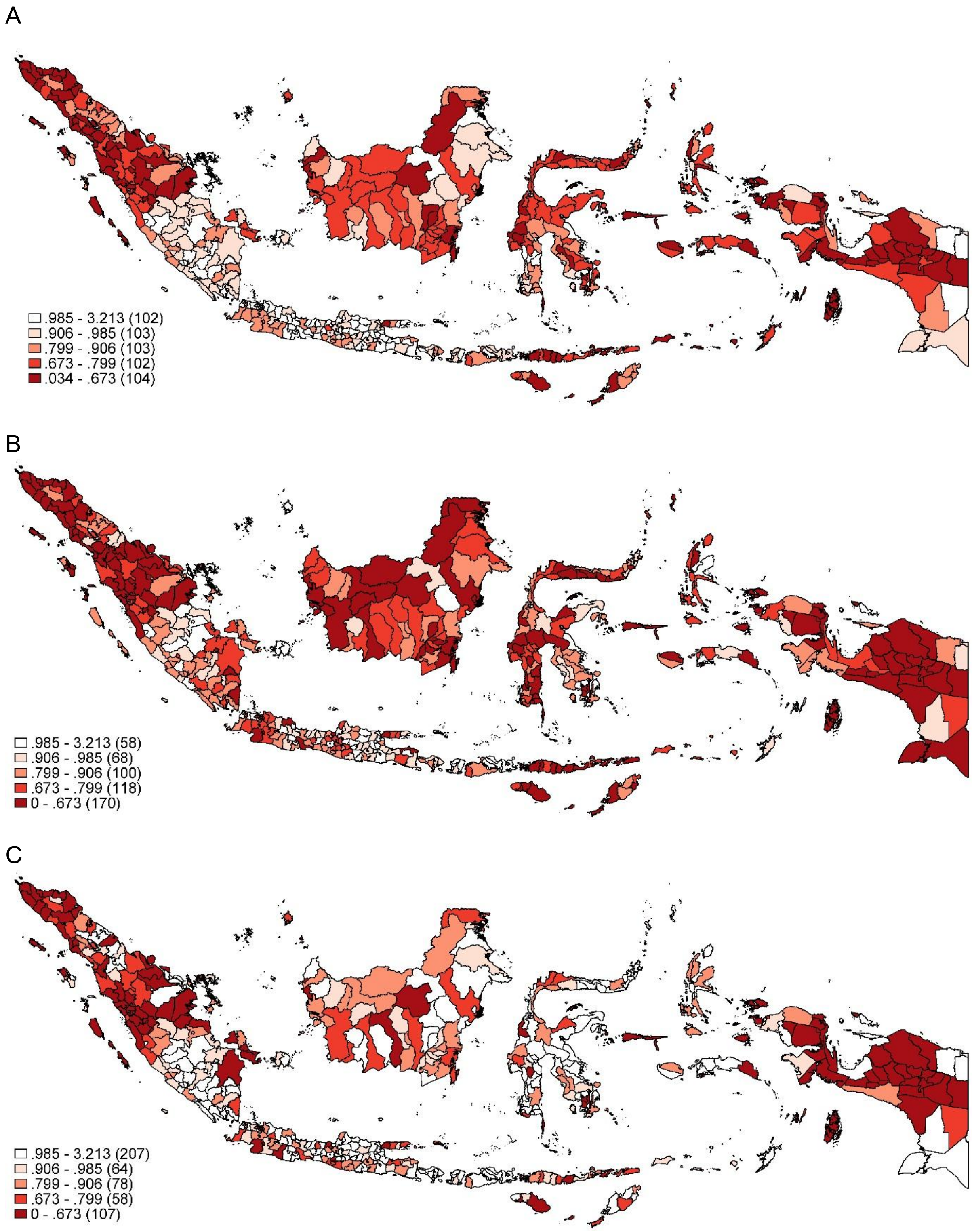


**Supplemental figure 1.** Geographical map of childhood immunization coverage across Indonesia during the pre-pandemic period (A), the first year of pandemic period (B), and the second year of pandemic period (C).

**Supplemental table 1**. Summary statistics of explanatory variables across 514 districts in Indonesia

| Variables | Median | Interquartile Range |
|---|---|---|
| **COVID-19 burden (2020–2021)** | | |
| Cumulative COVID-19 incidence rate (per 100,000 population) | 137 | (57 – 280) |
| Cumulative COVID-19 mortality rate (per 100,000 population) | 4 | (1 – 8) |
| **Public Health Development Indicators/PHDI (2018)** | | |
| Overall PHDI | 0.605 | (0.564 – 0.642) |
| Proportion of malnutrition in children under five | 18.5 | (14.6 – 23.6) |
| Proportion of stunting in children under five years old | 31.9 | (26.2 – 36.8) |
| Coverage of child weight monitoring | 78.6 | (67 – 87.4) |
| Coverage of neonatal visit | 85.6 | (76.3 – 91.8) |
| Proportion of health facility birth | 67 | (46.2 – 84.9) |
| Proportion of sub-district with adequate doctors | 8.3 | (0 – 20) |
| Proportion of village with adequate midwives | 31.7 | (6.9 – 81.5) |
| Proportion of village with adequate integrated community-based health post (posyandu) | 38.7 | (13.2 – 60.2) |
| Proportion of national health insurance ownership | 64.5 | (52 – 82.9) |
| **Human Development Indicators/HDI (2020)** | | |
| HDI | 69.3 | (66.4 – 73.0) |
| Domestic expenditure per capita (USD) | 620.6 | (522.1 – 713.2) |
| Average duration of formal education | 8.2 | (7.4 – 9.3) |
| Life expectancy at birth | 69.9 | (67.2 – 72.0) |
| **COVID-19 Vaccination Coverage** | | |
| $1^{st}$ dose coverage | 0.90 | (0.82 – 0.96) |
| $2^{nd}$ dose coverage | 0.63 | (0.52 – 0.74) |
